# Data-Driven Endocrine–Metabolic Phenotypes in Young Women With Polycystic Ovary Syndrome and Associations With Cardiometabolic Risk Markers

**DOI:** 10.64898/2026.02.25.26346893

**Authors:** Natalia Piórkowska, Kinga Nicigur, Marcin Leśniewski, Grzegorz Franik, Anna Bizoń

## Abstract

**Context:** Polycystic ovary syndrome (PCOS) is a heterogeneous endocrine disorder associated with reproductive dysfunction and long-term cardiometabolic risk. Traditional phenotype classifications based on diagnostic criteria may not fully capture the multidimensional biological variability underlying endocrine and metabolic risk profiles, particularly in young women.

**Objective:** To identify data-driven endocrine–metabolic phenotypes in young women with PCOS and evaluate their association with established cardiometabolic risk markers.

**Design and Setting:** Cross-sectional study conducted at a tertiary Gynecological Endocrinology Clinic in Poland between January 2018 and May 2025.

**Participants:** A total of 1300 young women diagnosed with PCOS according to Rotterdam criteria were included. The primary analytic cohort comprised 1032 participants aged 16–25 years with complete endocrine–metabolic biomarker data.

**Main Outcome Measures:** Endocrine–metabolic phenotypes were derived using principal component analysis followed by Gaussian mixture model clustering. Cardiometabolic risk endpoints included impaired glucose tolerance (2-hour plasma glucose during an oral glucose tolerance test ≥140 mg/dL), an atherogenic lipid profile (triglycerides (TG)/high-density lipoproteins (HDL-C) ratio >3.50), elevated non–HDL cholesterol (≥130 mg/dL), and a composite outcome of any abnormality.

**Results:** Principal component analysis retained 10 components explaining 81.9% of total variance. Unsupervised clustering identified two stable phenotypes (silhouette = 0.392; ARI = 0.842). Cluster 0 (n=954; 92.4%) represented a mixed endocrine–metabolic profile, whereas cluster 1 (n=78; 7.6%) was enriched for thyroid/autoimmune features, with higher anti–thyroid peroxidase antibody levels and higher thyroid-stimulating hormone. Cluster 1 showed a higher prevalence of an atherogenic lipid profile compared with cluster 0, while differences in glucose intolerance and non–HDL cholesterol were modest. Logistic regression analyses suggested phenotype-specific variation in cardiometabolic risk markers.

**Conclusions:** In a large cohort of young women with PCOS, data-driven analysis identified two reproducible endocrine–metabolic phenotypes, including a distinct thyroid/autoimmune-enriched subgroup. These findings highlight clinically relevant heterogeneity beyond traditional diagnostic phenotypes and support the potential value of integrated endocrine–metabolic profiling for early risk stratification in PCOS.

## Introduction

Polycystic ovary syndrome (PCOS) is one of the most common endocrine disorders in women of reproductive age and represents a leading cause of ovulatory dysfunction, hyperandrogenism, and reproductive morbidity (1). Beyond its gynecologic manifestations, PCOS is increasingly recognized as a systemic condition associated with metabolic abnormalities, including dyslipidemia, impaired glucose tolerance, and an elevated long-term risk of type 2 diabetes and cardiovascular disease (2,3). Importantly, PCOS is not a uniform disorder but rather a highly heterogeneous syndrome, with substantial variability in endocrine profiles, metabolic risk burden, and clinical trajectories across affected individuals (4). This heterogeneity has important implications for precision medicine, as women with PCOS may differ markedly in their trajectories of metabolic deterioration and long-term cardiometabolic risk (5).

Current clinical classification relies primarily on the Rotterdam diagnostic framework, which defines PCOS based on combinations of oligo-anovulation, hyperandrogenism, and polycystic ovarian morphology (6). Although useful for diagnosis, these traditional phenotypes (A–D) are limited in their ability to capture the full multidimensional biological diversity of PCOS (7,8). In particular, Rotterdam-based subtypes are criterion-driven rather than biology-driven and do not explicitly integrate broader endocrine axes or metabolic variation. As a result, clinically meaningful heterogeneity relevant to cardiometabolic risk stratification may remain underrecognized within standard diagnostic categories (9).

In recent years, there has been growing interest in applying data-driven subtyping approaches in endocrinology and metabolic medicine to refine disease classification beyond symptom-based frameworks (10). Unsupervised machine learning methods, including principal component analysis and clustering algorithms, have been used to identify biologically coherent endotypes in complex disorders, offering the potential to uncover latent phenotypic structure and improve individualized risk assessment (11,12). Within PCOS, emerging studies have suggested the presence of subgroups characterized by differential metabolic dysfunction, androgen excess, or inflammatory profiles; however, most work has focused on adult populations or has not comprehensively integrated multiple endocrine axes alongside cardiometabolic markers.

Notably, in young women with PCOS, early endocrine–metabolic phenotypes that jointly reflect gonadal, thyroid, and adrenal hormone regulation together with lipid and glucose metabolism remain poorly characterized. Crosstalk between ovarian steroidogenesis, thyroid function, adrenal hormone production, and metabolism signaling pathways is increasingly recognized as a key determinant of phenotypic expression in PCOS (13,14). Studying these relationships in adolescence and early adulthood is particularly relevant, as metabolic complications often evolve gradually and may not yet be clinically overt (15,16). Early identification of high-risk endocrine-metabolic phenotypes therefore represents a major clinical challenge, given that conventional screening strategies frequently rely on abnormalities that emerge later in the disease course (17).

Importantly, the application of data-driven phenotyping in the young PCOS population remains limited, despite the potential clinical value of identifying early endocrine-metabolic constellations that may precede overt cardiometabolic disease. Integrating biomarkers across multiple hormonal axes together with metabolic parameters may provide a more biologically grounded framework for risk stratification than traditional diagnostic classifications alone. Improved phenotypic resolution may facilitate earlier metabolic surveillance, refine risk prediction, and ultimately support more individualized management strategies in women with PCOS.

Therefore, the present study aimed to identify endocrine-metabolic phenotypes in a large cohort of young women with PCOS using an unsupervised, data-driven framework integrating biomarkers across key hormonal axes and cardiometabolic parameters.

We hypothesized that biologically coherent and clinically interpretable phenotypic clusters exist within PCOS and that these phenotypes differ in their association with established cardiometabolic risk markers, thereby offering potential utility for early cardiometabolic risk stratification.

## Methods

### Study design and participants

This cross-sectional study was conducted in the Gynecological Endocrinology Clinic of the Silesian Medical University in Katowice, Poland, between January 2018 and May 2025. The study included young women aged 16–25 years (mean age 20.9 years) who were hospitalized for diagnostic evaluation of PCOS.

PCOS was diagnosed according to the 2003 Rotterdam ESHRE/ASRM criteria, requiring the presence of at least two of the following features after exclusion of related disorders: (i) oligo-or anovulation, (ii) clinical and/or biochemical hyperandrogenism, and (iii) polycystic ovarian morphology (PCOM) on ultrasound (6).

### Assessment of polycystic ovarian morphology

PCOM was evaluated using transvaginal ultrasound (Voluson S8, GE Healthcare) with a 7.5-MHz vaginal probe. Ovaries were classified as polycystic based on the presence of ≥12 follicles measuring 2–8 mm in diameter and/or increased ovarian volume (>10 mL).

### Exclusion criteria

Women were excluded if alternative causes of hyperandrogenism or menstrual dysfunction were identified, including thyroid disease, hyperprolactinemia, nonclassic congenital adrenal hyperplasia, Cushing’s syndrome, or androgen-producing tumors. Women with diabetes mellitus or those receiving hormonal or metabolic therapies within the preceding 6 months were also excluded.

### Blood sampling procedures

Blood samples were obtained in the early follicular phase (cycle days 2–5) whenever possible, in the morning after an overnight fast of at least 8 hours. Metabolic assessment included fasting glucose and lipid profile. All participants underwent a standard 75-g oral glucose tolerance test (OGTT), with plasma glucose measured at baseline and 120 minutes.

### Ethics

The study protocol was approved by the Bioethical Committee of Wroclaw Medical University, Poland (KBN No. 254/2021) and was conducted in accordance with the Declaration of Helsinki.

### Input variables for phenotyping analysis

To identify endocrine–metabolic phenotypes within the PCOS cohort, a set of biologically relevant variables reflecting key hormonal axes and metabolic status was selected a priori. The feature set included markers of the gonadal axis (luteinizing hormone (LH), follicle stimulating hormone (FSH), LH/FSH ratio, total testosterone, sex-hormone binding globulin (SHBG)), thyroid function (thyroid-stimulating hormone (TSH), free thyroxine (FT4), anti-thyroid peroxidase (anti-TPO) antibodies), adrenal axis activity (dehydroepiandrosterone sulfate (DHEA-S), morning and evening cortisol, diurnal cortisol ratio), and metabolic parameters (total cholesterol (TCHOL), low-density lipoproteins cholesterol (LDL-C), high-density lipoproteins cholesterol (HDL-C) , triglycerides, fasting glucose, and 2-hour plasma glucose during an OGTT).

Derived indices, including triglyceride-to-HDL-C ratio and non–high-density lipoprotein cholesterol (non–HDL-C), were calculated as additional cardiometabolic risk markers. A complete list of input features and transformations is provided in Table S1.

Anti-TPO antibody concentrations were log-transformed prior to analysis due to their skewed distribution.

### Data preprocessing and construction of the analytic cohort

Raw endocrine and metabolic variables were harmonized prior to dimensionality reduction and clustering. Missingness patterns and feature distributions were assessed (Figure S1). Most core variables had low-to-moderate missingness (∼9–11%), whereas anti-TPO antibodies showed higher missingness (18.9%) and Anti-Müllerian hormone (AMH) was missing in 24.0% of participants (Table S2).

The primary phenotyping analysis was performed using a complete-case approach. Among 1300 women in the raw dataset, 1032 participants (79.4%) had complete data available across the core endocrine–metabolic feature set and were retained for principal component analysis (PCA) and clustering, while 268 women were excluded due to missingness (Figure S2).

The main contributors to exclusion were missing anti-TPO antibody measurements (245 excluded participants), followed by missing OGTT-derived glucose values at 120 minutes (138 excluded participants) and fasting glucose (137 excluded participants) (Table S2).

All continuous variables were standardized using z-score normalization. Winsorization at the 1st–99th percentile was pre-specified as a sensitivity procedure but was not applied in the primary analysis. All analyses were performed with a fixed random seed (42).

### Dimensionality reduction

To reduce collinearity among endocrine–metabolic biomarkers and obtain a lower-dimensional representation for clustering, PCA was applied to the standardized complete-case feature matrix.

The number of retained components was determined using scree plot inspection and cumulative explained variance (Figure S3A). The first 10 principal components accounted for 81.9% of total variance and were carried forward into clustering.

Component loadings supported clinical interpretability. PC1 (19.9% variance) was driven by lipid-related cardiometabolic markers (non–HDL-C, triglycerides (TG), TG/HDL-C ratio, LDL-C), consistent with a dyslipidemic–metabolic axis. PC3 reflected a gonadotropin-related reproductive axis, whereas PC5 was characterized by thyroid/autoimmune features. Full PCA loadings are provided in Figure S3B and Table S3.

### Clustering analysis and validation

Unsupervised clustering was performed in the reduced PCA space using Gaussian mixture models (GMM), which allow probabilistic class membership and flexible cluster geometry.

The optimal number of clusters was evaluated across K = 2–10 using silhouette coefficient, cluster stability under subsampling, and information criteria (AIC/BIC) (Figure S4A–C). To avoid clinically non-meaningful fragmentation, a minimum cluster size constraint was imposed (minimum cluster fraction ≥5%).

The final selected solution was a 2-cluster GMM model (silhouette = 0.392; BIC = 26429.6; AIC = 25782.5). Models with K ≥3 were rejected due to formation of unstable low-prevalence clusters (Figure S4D).

Cluster stability was high (mean adjusted Rand index [ARI] = 0.842). Membership certainty was strong, with a mean maximum posterior probability of 0.989 (Table S4).

In the final analytic cohort (n = 1032), cluster 0 included 954 women (92.4%), whereas cluster 1 included 78 women (7.6%). Cluster visualization in PCA space and UMAP projections are shown in Figure S5.

### Cardiometabolic risk outcomes

Cardiometabolic risk markers were defined a priori. Impaired glucose tolerance was defined as 2-hour OGTT glucose ≥140 mg/dL (ADA criterion). Additional lipid-based endpoints included TG/HDL-C ratio >3.50 and elevated non–HDL-C ≥130 mg/dL. A composite outcome (“any cardiometabolic abnormality”) was defined as the presence of ≥1 endpoint. Endpoint definitions are summarized in Table S5.

### Statistical analysis

Continuous variables were summarized as medians with interquartile ranges and compared between clusters using Kruskal–Wallis tests. Categorical risk endpoints were compared using chi-square tests with Yates correction or Fisher’s exact test when expected counts were small.

Multiple testing correction across endpoints was performed using the Benjamini–Hochberg false discovery rate procedure.

Logistic regression models were fitted to evaluate associations between cluster membership and cardiometabolic risk endpoints, using cluster 0 as the reference group. Models were estimated with robust standard errors and restricted to endpoints with at least 10 events. Age (available for all participants; range 16–25 years) was included as an adjustment covariate in primary analyses. Full regression outputs are provided in Table S6 and visualized in Figure S6.

### Post-hoc biological validation

AMH was not included in the primary clustering feature set and was analyzed post hoc as an external biological validator. AMH concentrations were available for 970 of 1032 participants (94.0%). Median AMH levels were similar across clusters, with no statistically significant difference observed (p=0.18) (Figure S7, Table S7).

### Software and reproducibility

All preprocessing, dimensionality reduction, clustering, and regression analyses were performed in Python using a fixed random seed (42). Statistical modeling was conducted using the statsmodels package (version 0.14.6).

## Results

### Cohort characteristics

A total of 1300 young women with PCOS were initially included in the raw dataset. After preprocessing and complete-case selection, 1032 participants (79.4%) with complete endocrine–metabolic data were retained for PCA and clustering analyses, while 268 were excluded due to missingness in one or more core variables.

The analytic cohort was aged 16–25 years (mean age 20.9 years). Baseline endocrine and metabolic characteristics of the overall cohort are summarized in Table 1.

**Table 1.**
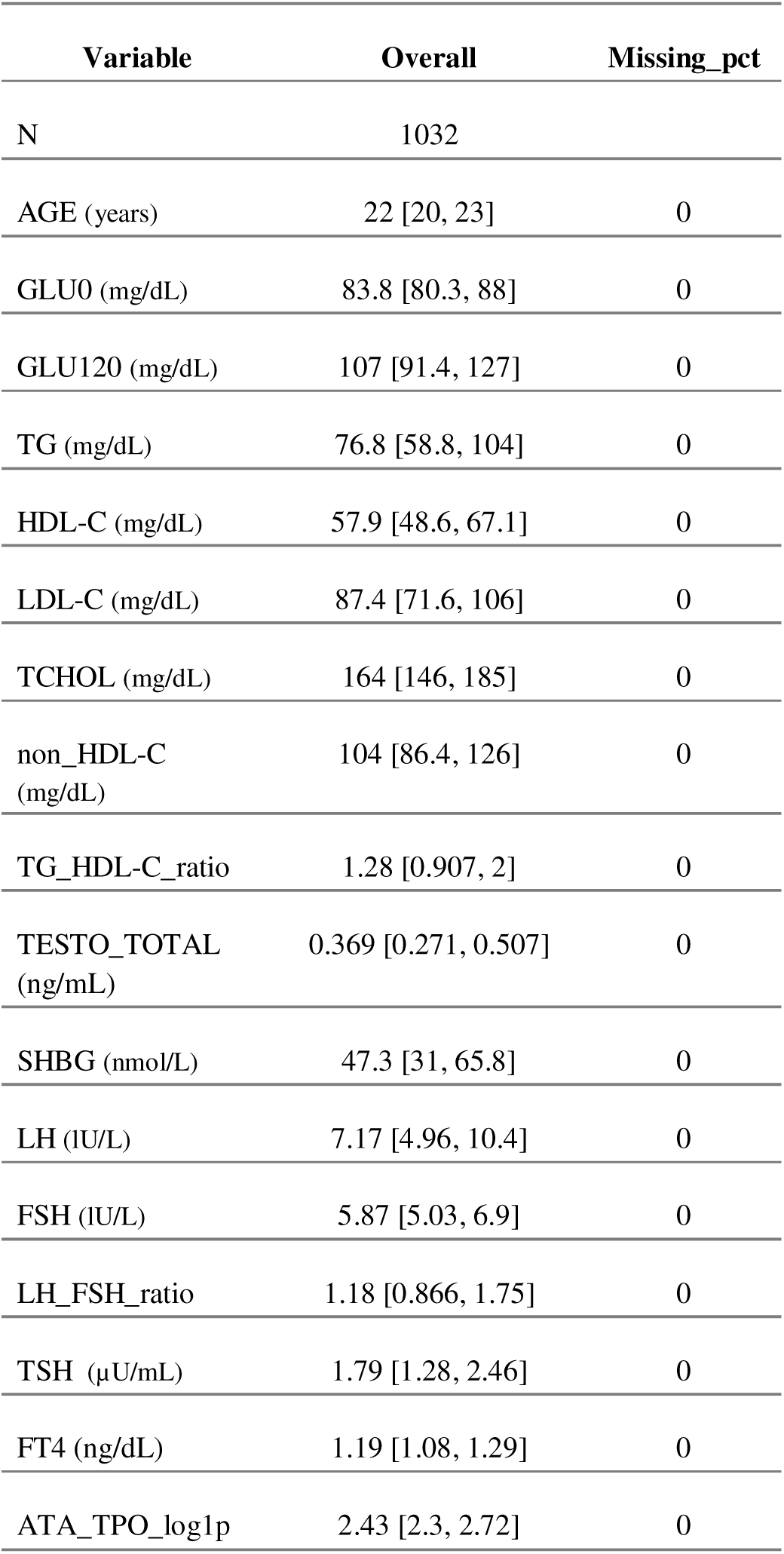
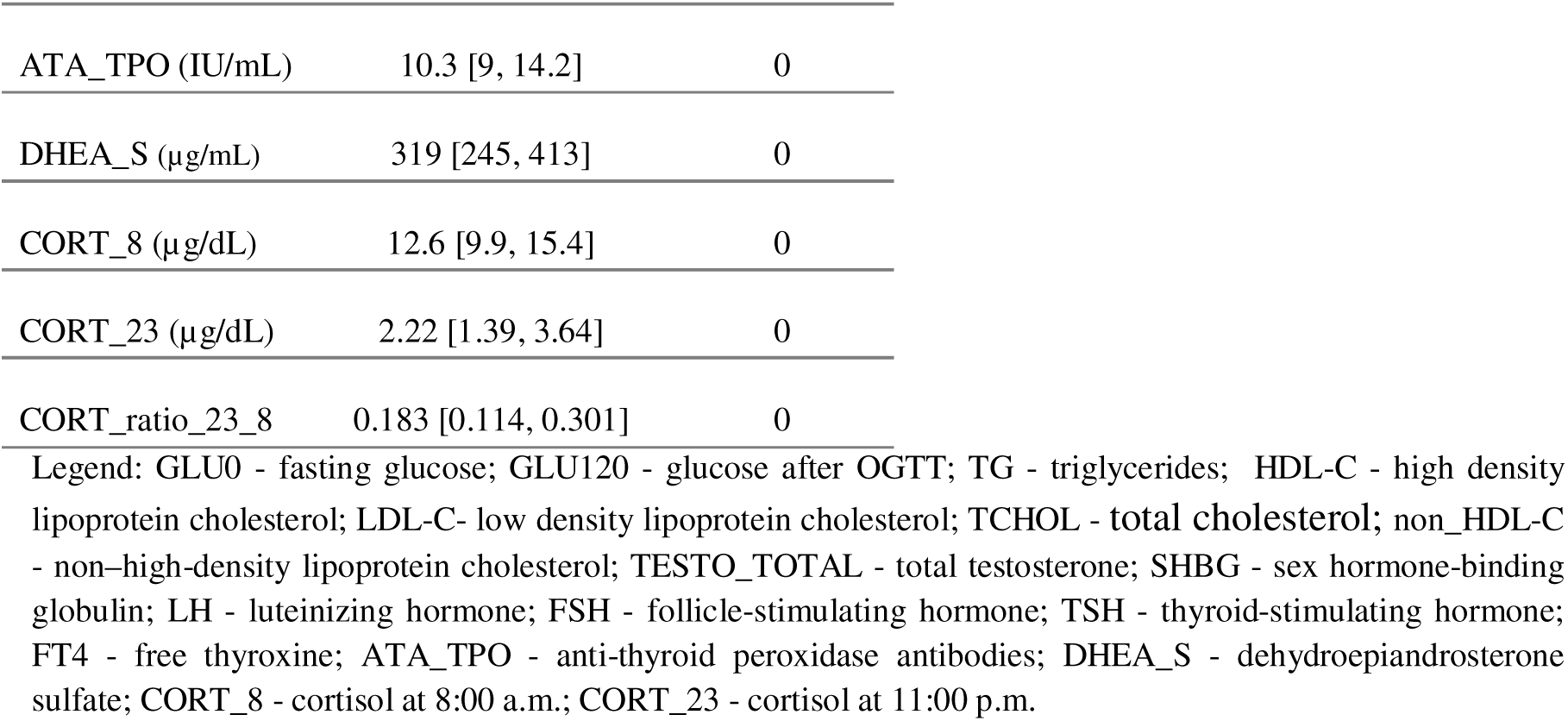
Baseline endocrine and metabolic characteristics of the analytic PCOS cohort (n=1032). Values are presented as median (interquartile range) unless otherwise indicated.

| Variable | Overall | Missing_pct |
| --- | --- | --- |
| N | 1032 |  |
| AGE (years) | 22 [20, 23] | 0 |
| GLU0 (mg/dL) | 83.8 [80.3, 88] | 0 |
| GLU120 (mg/dL) | 107 [91.4, 127] | 0 |
| TG (mg/dL) | 76.8 [58.8, 104] | 0 |
| HDL-C (mg/dL) | 57.9 [48.6, 67.1] | 0 |
| LDL-C (mg/dL) | 87.4 [71.6, 106] | 0 |
| TCHOL (mg/dL) | 164 [146, 185] | 0 |
| non_HDL-C<br>(mg/dL) | 104 [86.4, 126] | 0 |
| TG_HDL-C_ratio | 1.28 [0.907, 2] | 0 |
| TESTO_TOTAL<br>(ng/mL) | 0.369 [0.271, 0.507] | 0 |
| SHBG (nmol/L) | 47.3 [31, 65.8] | 0 |
| LH (IU/L) | 7.17 [4.96, 10.4] | 0 |
| FSH (IU/L) | 5.87 [5.03, 6.9] | 0 |
| LH_FSH_ratio | 1.18 [0.866, 1.75] | 0 |
| TSH (μU/mL) | 1.79 [1.28, 2.46] | 0 |
| FT4 (ng/dL) | 1.19 [1.08, 1.29] | 0 |
| ATA_TPO_log1p | 2.43 [2.3, 2.72] | 0 |
| ATA_TPO (IU/mL) | 10.3 [9, 14.2] | 0 |
| DHEA_S (μg/mL) | 319 [245, 413] | 0 |
| CORT_8 (μg/dL) | 12.6 [9.9, 15.4] | 0 |
| CORT_23 (μg/dL) | 2.22 [1.39, 3.64] | 0 |
| CORT_ratio_23_8 | 0.183 [0.114, 0.301] | 0 |

### Dimensionality reduction by PCA

Principal component analysis was applied to the standardized endocrine–metabolic feature matrix to reduce collinearity and derive a lower-dimensional representation for clustering.

The first 10 principal components explained 81.9% of total variance and were retained for downstream modeling. PC1 accounted for 19.9% of variance and was primarily driven by cardiometabolic lipid markers (non–HDL-C, TG, TG/HDL-C ratio, LDL-C), consistent with a dyslipidemic–metabolic axis. PC3 reflected a gonadotropin-related reproductive endocrine axis, whereas PC5 was characterized by thyroid/autoimmune features (anti-TPO antibodies and TSH).

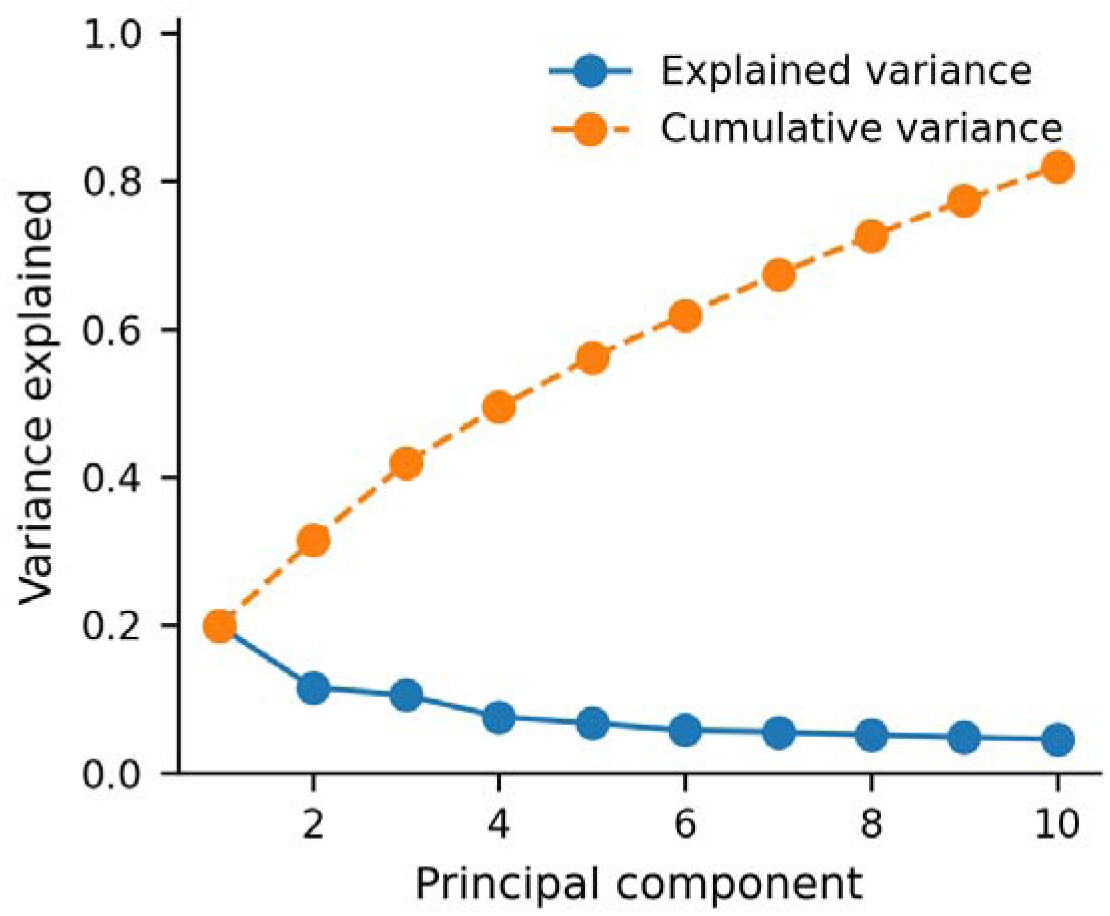

A scree plot and PCA loading heatmap are shown in Figure 1 (see also Table S3).

**Figure 1.**
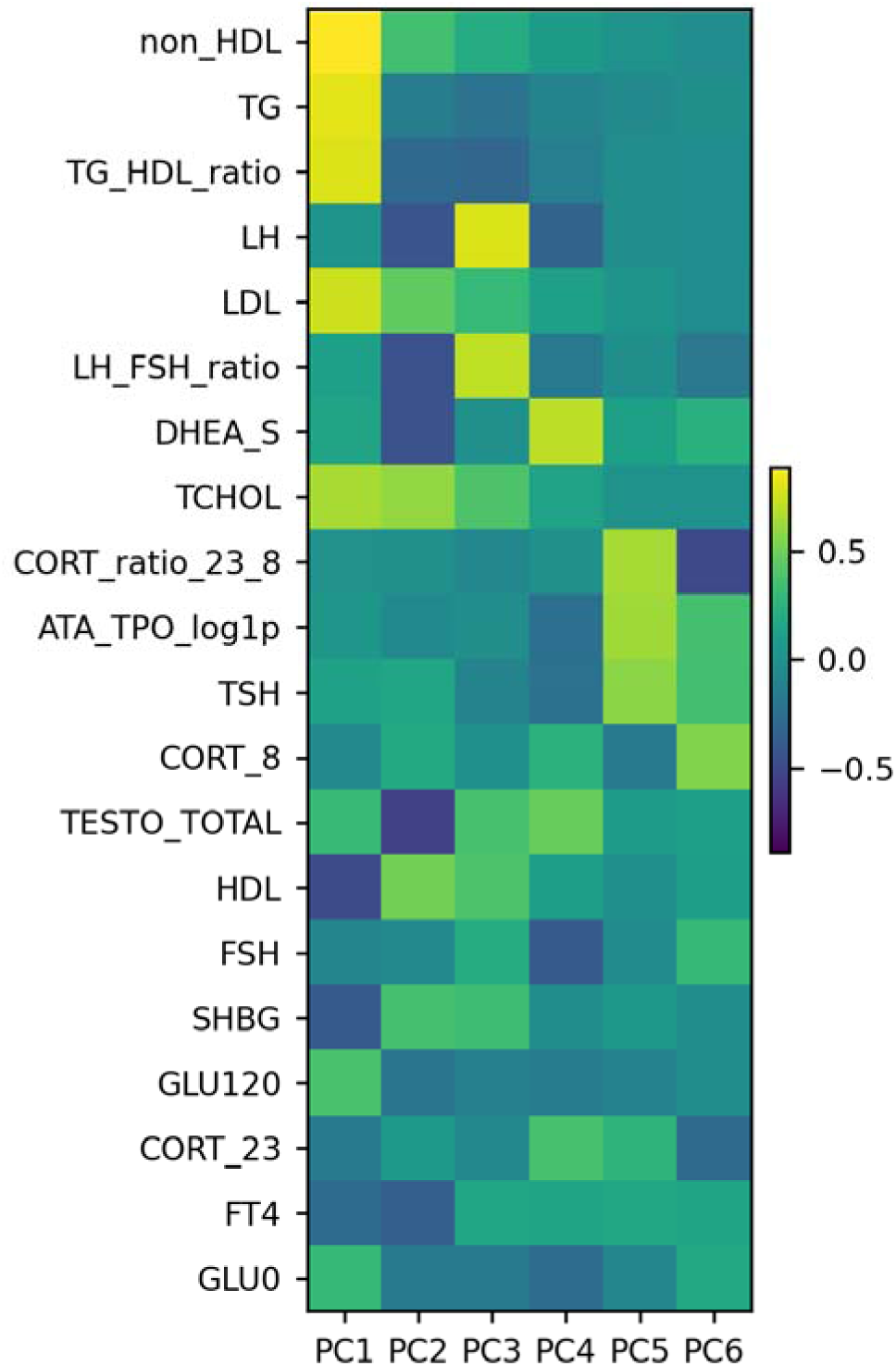
Scree plot of explained variance (A) and heatmap of principal component loadings (B). Panel A shows the proportion of variance explained by each principal component and the cumulative explained variance. Panel B displays features ranked by absolute loading across the leading principal components derived from z-standardized core variables in the complete-case analytic cohort. Legend: GLU0 - fasting glucose; GLU120 - glucose after OGTT; TG - triglycerides; HDL-C - high density lipoprotein cholesterol; LDL-C- low density lipoprotein cholesterol; TCHOL - total cholesterol; non_HDL-C - non–high-density lipoprotein cholesterol; TESTO_TOTAL - total testosterone; SHBG - sex hormone-binding globulin; LH - luteinizing hormone; FSH - follicle-stimulating hormone; TSH - thyroid-stimulating hormone; FT4 - free thyroxine; ATA_TPO - anti-thyroid peroxidase antibodies; DHEA_S - dehydroepiandrosterone sulfate; CORT_8 - cortisol at 8:00 a.m.; CORT_23 - cortisol at 11:00 p.m.

### Clustering analysis and stability

Unsupervised clustering in PCA space was performed using Gaussian mixture models. The optimal number of clusters was evaluated across K=2–10 using silhouette coefficient, AIC/BIC, and stability metrics.

A 2-cluster solution was selected as the most stable and clinically interpretable model (silhouette = 0.392; BIC = 26429.6; AIC = 25782.5). Models with K≥3 were rejected due to the emergence of unstable low-prevalence clusters below the minimum size threshold.

Cluster stability was high, with a mean adjusted Rand index (ARI) of 0.842. Membership certainty was strong (mean maximum posterior probability = 0.989).

Cluster visualization in PCA space is provided in Figure 2.

**Figure 2.**
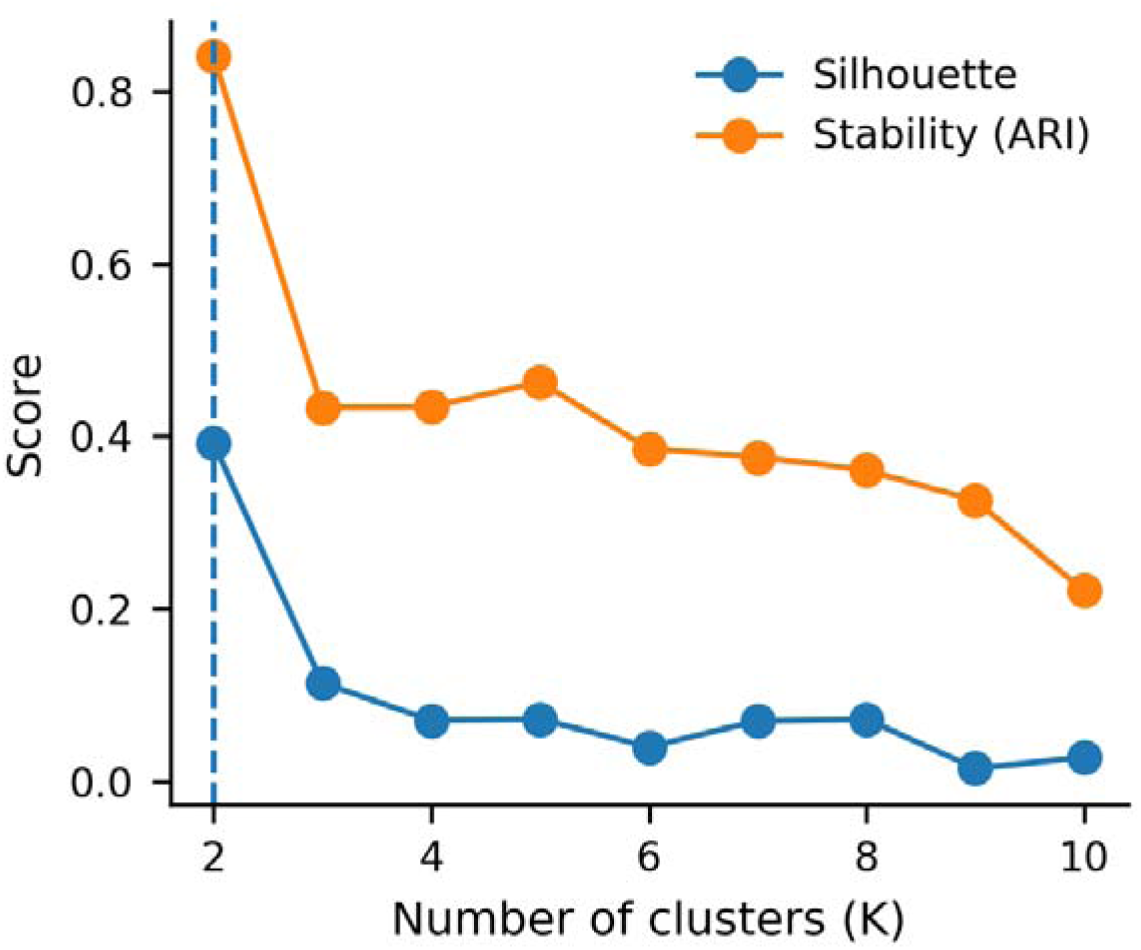
Model selection metrics (A) and PCA visualization of the final clustering solution (B). Panel A shows silhouette coefficient and cluster stability (mean adjusted Rand index) across K = 2–10 Gaussian mixture models, with the selected two-cluster solution indicated. Panel B displays the distribution of participants in PCA space (PC1 vs PC2), colored according to cluster assignment from the final two-cluster model.

### Endocrine–metabolic phenotypes of PCOS

The final model identified two endocrine–metabolic phenotypes within the PCOS cohort.

Cluster 0 represented the predominant phenotype (n=954; 92.4%), characterized by a mixed endocrine–metabolic profile without a single dominant axis.

Cluster 1 comprises a smaller subgroup (n=78; 7.6%) enriched for thyroid/autoimmune features, with higher anti-TPO antibody levels and higher TSH concentrations.

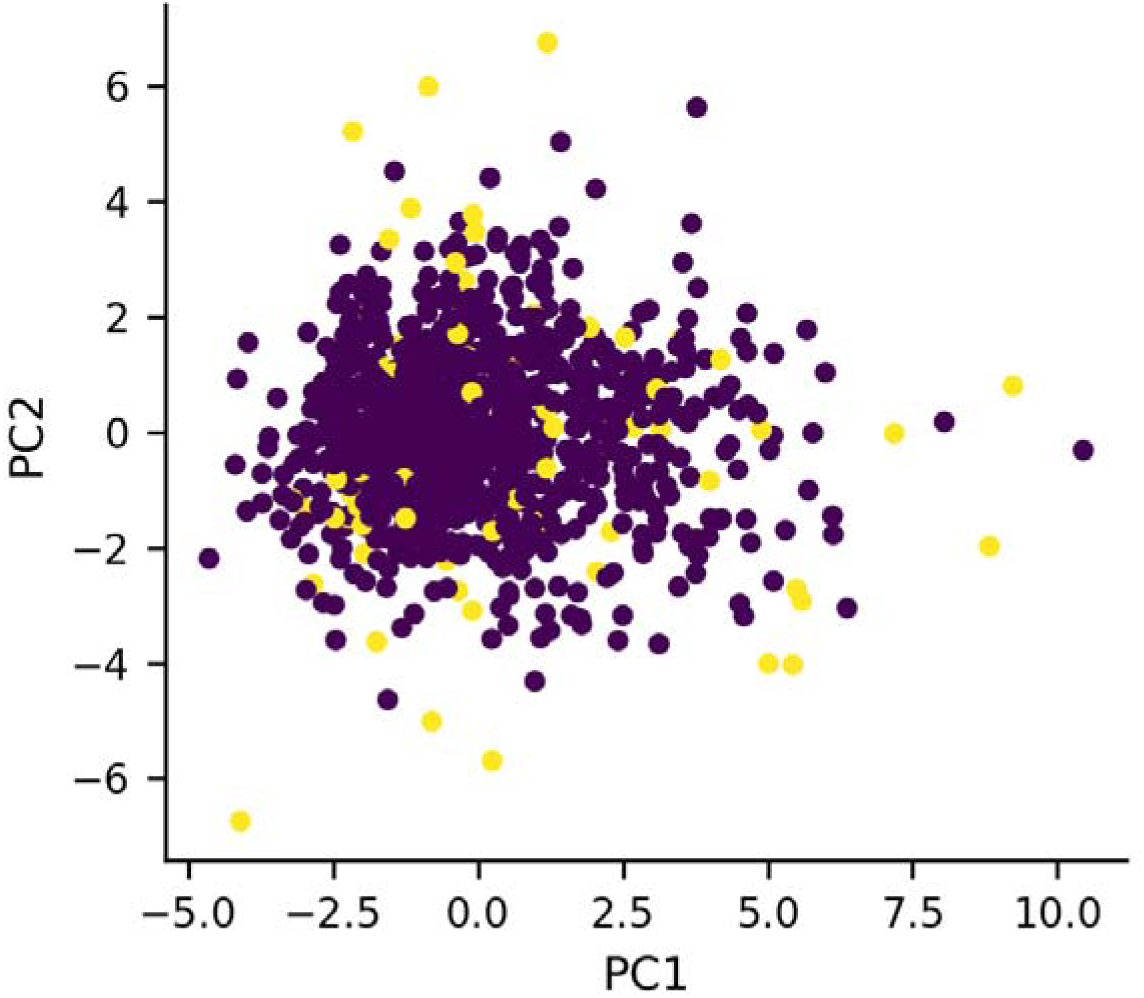

Median values and interquartile ranges for all hormonal and metabolic variables across clusters are presented in Table 2, while a standardized heatmap of cluster profiles is shown in Figure 3.

**Figure 3.**
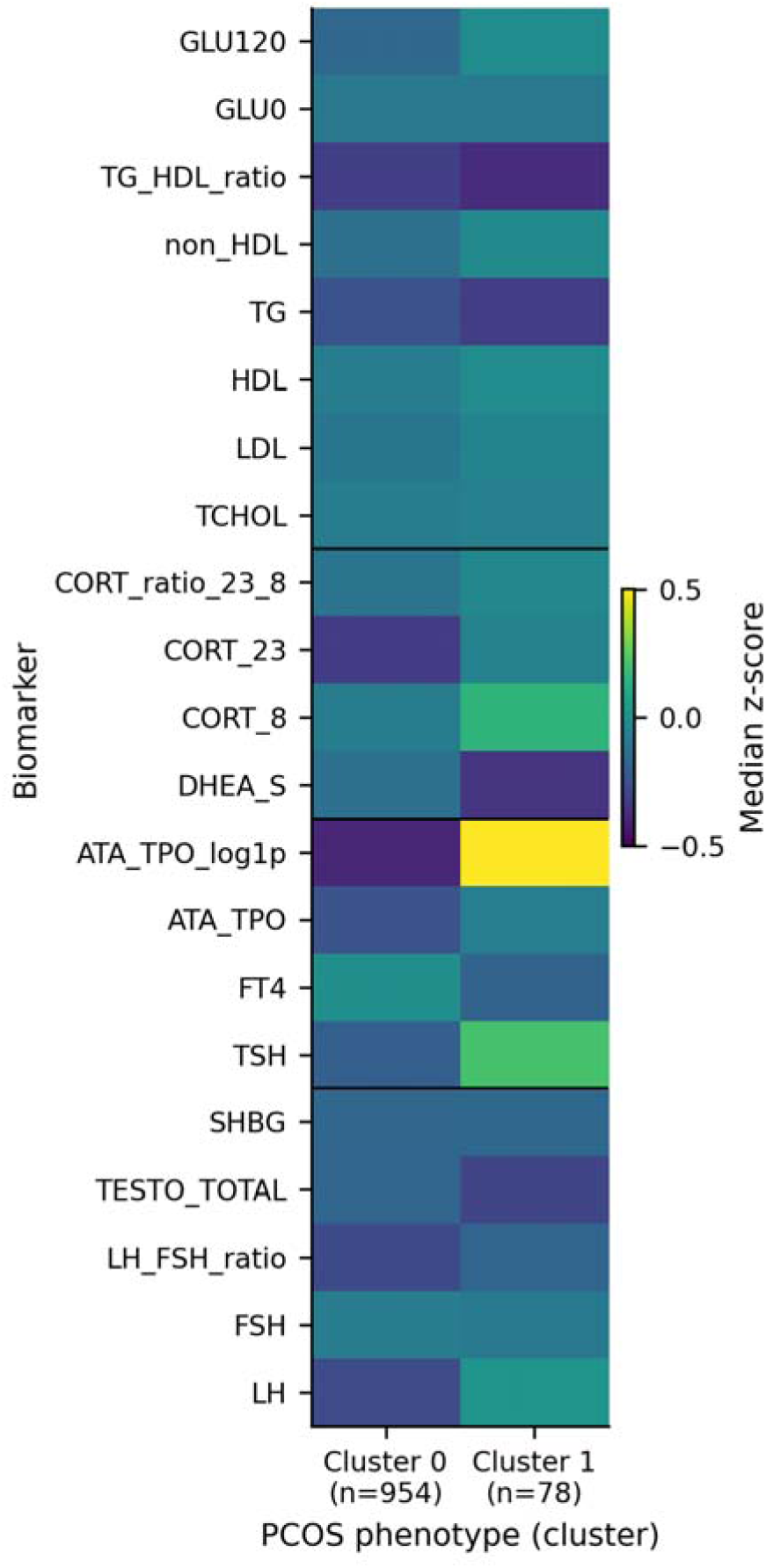
Heatmap of standardized endocrine and metabolic biomarkers across identified PCOS phenotypes. Values represent median z-scores within each cluster. Colors represent median z-scores within each cluster, centered at zero (white), with red indicating higher and blue lower standardized values relative to the overall cohort. Legend: GLU0 - fasting glucose; GLU120 - glucose after OGTT; TG - triglycerides; HDL-C - high density lipoprotein cholesterol; LDL-C- low density lipoprotein cholesterol; TCHOL - total cholesterol; non_HDL-C - non–high-density lipoprotein cholesterol; TESTO_TOTAL - total testosterone; SHBG - sex hormone-binding globulin; LH - luteinizing hormone; FSH - follicle-stimulating hormone; TSH - thyroid-stimulating hormone; FT4 - free thyroxine; ATA_TPO - anti-thyroid peroxidase antibodies; DHEA_S - dehydroepiandrosterone sulfate; CORT_8 - cortisol at 8:00 a.m.; CORT_23 - cortisol at 11:00 p.m.

**Table 2.** Endocrine–metabolic profiles of PCOS phenotypes identified by Gaussian mixture model clustering. Values are presented as median (IQR). P-values were adjusted using the Benjamini–Hochberg false discovery rate procedure.

|  |  |  |  |
| --- | --- | --- | --- |
| GLU0 (mg/dL) | 83.8 [80.3, 87.9] | 83.8 [79.6, 88.7] | 0.887 |
| GLU120 (mg/dL) | 107 [91.4, 127] | 112 [93.2, 138] | 0.501 |
| TG (mg/dL) | 77 [59.1, 104] | 73.8 [57.8, 108] | 0.937 |
| HDL-C (mg/dL) | 57.7 [49, 67.5] | 58.5 [47.1, 64.9] | 0.887 |
| LDL-C (mg/dL) | 87.3 [71.8, 105] | 88.9 [70.6, 112] | 0.923 |
| TCHOL (mg/dL) | 164 [146, 184] | 164 [143, 193] | 0.887 |
| non_HDL-C (mg/dL) | 104 [86.5, 125] | 107 [83.2, 131] | 0.887 |
| TG_HDL-C_ratio | 1.29 [0.906, 2] | 1.21 [0.926, 2.01] | 0.887 |
| TESTO_TOTAL<br>(ng/mL) | 0.37 [0.272, 0.512] | 0.347 [0.235, 0.466] | 0.501 |
| SHBG (nmol/L) | 47.3 [31, 65.8] | 47.5 [31.3, 68] | 0.937 |
| LH (IU/L) | 7.05 [4.96, 10.2] | 8.65 [5.56, 14.6] | 0.027 |
| FSH (IU/L) | 5.88 [5.05, 6.9] | 5.78 [4.65, 6.87] | 0.501 |
| LH_FSH_ratio | 1.17 [0.865, 1.7] | 1.26 [0.871, 2.19] | 0.450 |
| TSH (μU/mL) | 1.77 [1.27, 2.42] | 2.24 [1.4, 3.83] | 0.001 |
| FT4 (ng/dL) | 1.19 [1.09, 1.29] | 1.16 [1.04, 1.33] | 0.887 |
| ATA_TPO_log1p | 2.4 [2.3, 2.66] | 3.04 [2.55, 5.54] | 0.000 |
| ATA_TPO (IU/mL) | 10 [9, 13.3] | 20.1 [11.8, 255] | 0.000 |
| DHEA_S (μg/mL) | 321 [245, 416] | 290 [245, 358] | 0.077 |
| CORT_8 (μg/dL) | 12.5 [9.91, 15.4] | 13.4 [9.84, 16.3] | 0.887 |
| CORT_23 (μg/dL) | 2.2 [1.38, 3.58] | 2.87 [1.6, 10.8] | 0.003 |
| CORT_ratio_23_8 | 0.18 [0.114, 0.292] | 0.245 [0.121, 0.822] | 0.003 |

Given the dominant thyroid-related signal, cluster 1 was conservatively referred to as a “thyroid/autoimmune-enriched phenotype,” whereas cluster 0 was considered the reference mixed phenotype.

### Cardiometabolic risk across phenotypes

Cardiometabolic risk endpoints were compared across clusters, including impaired glucose tolerance (2-hour OGTT glucose ≥140 mg/dL), TG/HDL-C ratio >3.50, elevated non–HDL-C≥130 mg/dL, and a composite endpoint defined as the presence of any abnormality.

The prevalence of these endpoints by cluster is summarized in Table 3. Cluster 1 showed a higher frequency of an atherogenic lipid profile defined by TG/HDL-C >3.50 (χ² p=0.024; BH-FDR q=0.097), whereas no statistically significant differences were observed for glucose intolerance or elevated non–HDL-C.

**Table 3.** Prevalence of predefined cardiometabolic risk markers across endocrine–metabolic PCOS phenotypes. Odds ratios are reported relative to cluster 0 as the reference group.

| endpoint | Cluster 0<br>n(%) | Cluster 1<br>n(%) | OR(95% CI) | p-value | q-value (BH-FDR) |
| --- | --- | --- | --- | --- | --- |
| risk_glu120_ge_thr | 140 (14.7%) | 16 (20.5%) | 1.50 (0.84–2.67) | 0.169 | 0.210 |
| risk_nonhdl_ge_thr | 199 (20.9%) | 21 (26.9%) | 1.40 (0.83–2.36) | 0.210 | 0.210 |
| risk_tghdl_gt_thr | 58 (6.1%) | 10 (12.8%) | 2.27 (1.11–4.64) | 0.024 | 0.098 |
| risk_any_endpoint | 307 (32.2%) | 33 (42.3%) | 1.55 (0.97–2.47) | 0.069 | 0.138 |

Odds ratios were estimated using logistic regression models adjusted for age.

Logistic regression analyses estimating odds ratios for cardiometabolic risk by phenotype are presented in Figure 4 and Table S6.

**Figure 4.**
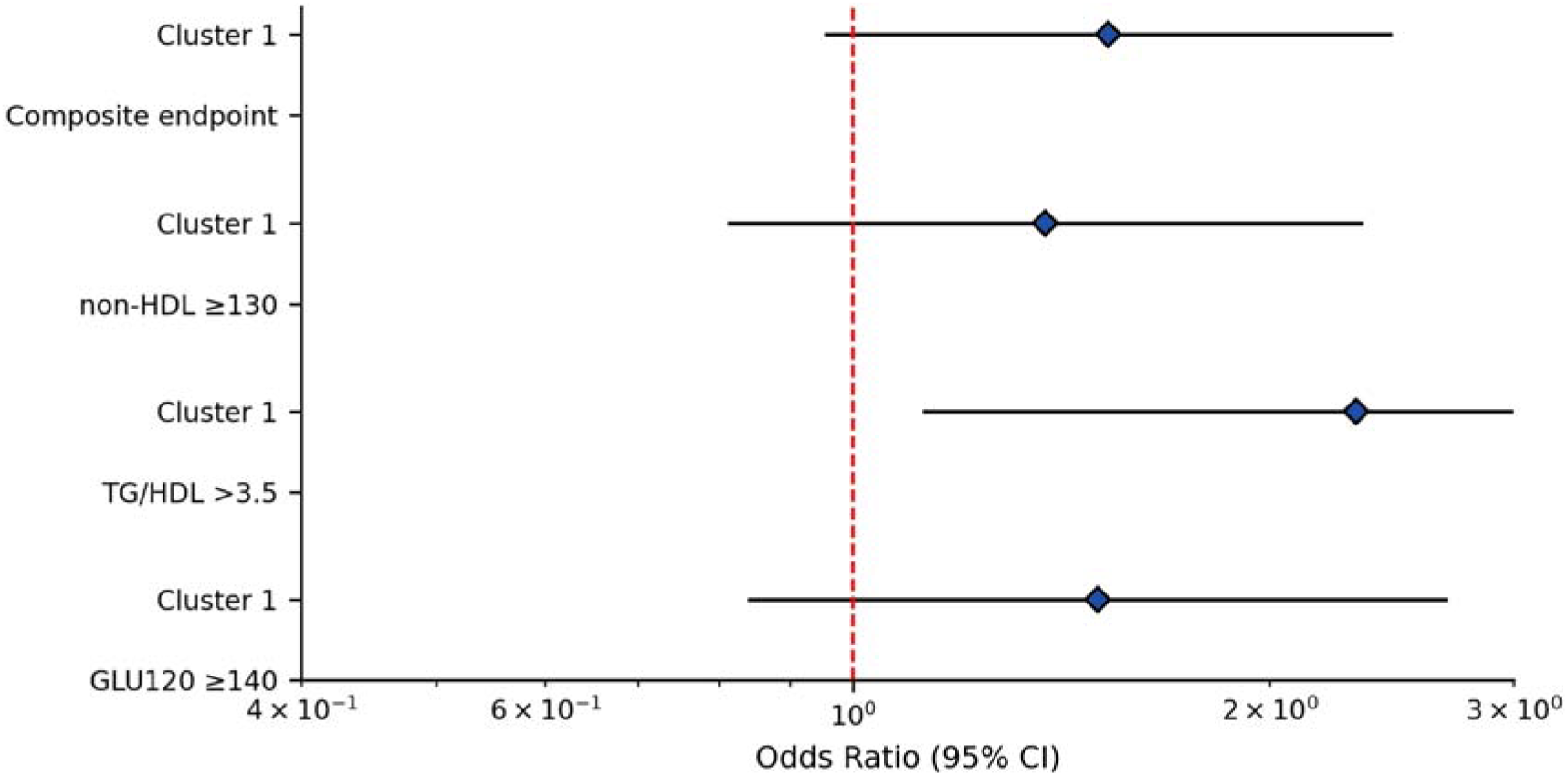
Age-adjusted odds ratios (ORs) and 95% confidence intervals for predefined cardiometabolic risk endpoints according to endocrine–metabolic PCOS phenotype (cluster 1 vs cluster 0 reference), estimated using logistic regression with robust standard errors.

### Post-hoc biological validation with AMH

As an external biological validator, AMH was analyzed post hoc and was not included in the clustering feature set. AMH concentrations were available for 970 participants (94.0%) and did not differ significantly between clusters (p=0.18), supporting that cluster separation was not driven by ovarian reserve markers.

### Discussion Main findings

In this cross-sectional, data-driven phenotyping study of young women with PCOS, we identified two stable endocrine–metabolic phenotypes by integrating reproductive, thyroid, adrenal, and metabolic biomarkers within an unsupervised clustering framework. The final Gaussian mixture model solution showed high robustness, supported by strong stability metrics and membership certainty, suggesting that the derived subgroups reflect reproducible biological structure rather than methodological noise. Although the silhouette coefficient indicated moderate separation, the high cluster stability (ARI = 0.842) and strong posterior membership probabilities support the robustness and reproducibility of the identified phenotypes.

The predominant phenotype (cluster 0) encompassed the majority of participants and demonstrated a broadly mixed endocrine–metabolic profile. In contrast, a smaller but distinct subgroup (cluster 1) was characterized by a prominent thyroid/autoimmune signal, with higher anti-TPO antibody levels and higher TSH concentrations. Cardiometabolic differentiation between clusters was modest in this young cohort, although an atherogenic lipid pattern reflected by TG/HDL-C ratio emerged as the clearest metabolic risk-related marker.

The metabolic relevance of TG and HDL-C in PCOS is further supported by our previous study (18), in which we demonstrated significant associations between alterations in TG and HDL-C concentrations and changes in SHBG levels and the free androgen index (FAI). Higher quartiles of TG concentration were associated with increased odds of decreased SHBG and elevated FAI, whereas higher HDL-C levels were linked to a lower probability of androgen-related disturbances. These findings reinforce the close interplay between lipid metabolism and hormonal dysregulation in PCOS.

Together, these findings highlight that even in early adulthood, PCOS heterogeneity extends beyond classical reproductive endocrine features and may incorporate clinically relevant thyroid–autoimmune variation.

### Biological interpretation of endocrine–metabolic phenotypes

PCOS is increasingly understood as a heterogeneous syndrome arising from complex interactions between hypothalamic–pituitary–ovarian dysfunction, adrenal androgen activity, metabolic regulation, and comorbid endocrine conditions. This conceptual shift has been emphasized in major pathophysiological reviews, which describe PCOS as a systemic endocrine-metabolic disorder involving partially independent but interacting axes, including ovarian steroidogenesis, insulin signaling, adrenal androgen production, and neuroendocrine regulation (19,20).

The thyroid/autoimmune-enriched phenotype observed in our cohort may reflect the recognized overlap between PCOS and autoimmune thyroid disease, which has been reported more frequently in women with PCOS than in the general population. Several meta-analyses have demonstrated a significantly higher prevalence of autoimmune thyroiditis and anti-TPO positivity in women with PCOS compared with controls (21,22), suggesting a non-random coexistence of these conditions.

Elevated anti-TPO antibodies together with higher TSH may indicate a subgroup in whom autoimmune thyroid processes contribute to systemic endocrine variability. Thyroid hormones influence lipid metabolism, hepatic lipoprotein turnover, and insulin sensitivity, which may partly explain the subtle lipid signal observed in this subgroup. Although overt metabolic differences were limited in our analysis, thyroid dysfunction and autoimmunity may still modulate lipid metabolism and cardiometabolic trajectories over time, potentially representing an early risk context rather than an established metabolic phenotype.

Importantly, we did not observe strong cluster separation driven purely by hyperandrogenism or severe metabolic impairment. This may partly reflect the young age and early disease stage of the cohort, in which cardiometabolic complications may not yet be fully manifest. Thus, endocrine heterogeneity may precede the development of more pronounced metabolic morbidity later in life.

### Comparison with existing literature

Traditional PCOS phenotyping based on Rotterdam criteria (phenotypes A–D) provides a useful diagnostic framework but does not fully capture multidimensional biological variation across endocrine axes. In recent years, unsupervised machine learning approaches have been increasingly applied to refine PCOS subtyping and identify biologically meaningful endotypes, often highlighting subgroups with metabolic dysfunction, hyperandrogenic severity, or inflammatory profiles. Genetic clustering analyses have identified at least two biologically distinct PCOS subtypes (8), including a reproductive and a metabolic subtype, supporting the concept that conventional diagnostic phenotypes incompletely capture underlying pathobiology. Unlike prior studies primarily distinguishing reproductive versus metabolic subtypes, our findings suggest that thyroid autoimmunity may represent an additional dimension of endocrine stratification in early PCOS.

Our results are consistent with this emerging paradigm but extend it in two important ways. First, the study focuses on a uniquely young population, providing insight into endocrine–metabolic heterogeneity at an early stage of reproductive life. Second, the identification of a thyroid/autoimmune-enriched subgroup underscores that PCOS subtyping may benefit from incorporating broader endocrine comorbidity markers beyond gonadal hormones and metabolic traits alone.

The presence of a relatively small but distinct autoimmune-related phenotype suggests that thyroid status may represent an additional dimension of PCOS heterogeneity that is not captured by classical diagnostic phenotypes.

### Clinical implications

From a clinical perspective, the identification of endocrine–metabolic phenotypes supports future efforts toward risk stratification and personalized screening in PCOS. Although cardiometabolic differences were modest in this cohort, the enrichment of thyroid autoimmunity suggests that a subset of young women with PCOS may benefit from more systematic thyroid evaluation and longitudinal follow-up.

Moreover, the observed lipid-related signal in the thyroid-enriched phenotype raises the possibility that endocrine comorbidities may interact with early cardiometabolic risk markers. As PCOS is associated with long-term metabolic and cardiovascular morbidity, early phenotypic stratification could eventually inform targeted prevention strategies, including more intensive metabolic monitoring in biologically higher-risk subgroups. The TG/HDL-C ratio has been proposed as a surrogate marker of atherogenic dyslipidemia and insulin resistance, particularly in young populations (23), and has been associated with cardiometabolic risk in PCOS (24). Even subtle lipid shifts in early adulthood may reflect future cardiometabolic divergence.

Longitudinal data indicate that cardiometabolic abnormalities in PCOS often evolve progressively across the lifespan (25), supporting the interpretation that endocrine divergence may precede overt metabolic morbidity.

More broadly, our findings reinforce the concept that PCOS classification may evolve from purely diagnostic criteria toward multidimensional endocrine–metabolic profiling with translational relevance. The movement toward biologically informed subtyping parallels precision medicine approaches applied in other complex metabolic disorders, including type 2 diabetes (26).

### Strengths and limitations

Several limitations should be acknowledged. First, the cross-sectional design precludes causal inference and limits the ability to determine whether the identified phenotypes predict future cardiometabolic outcomes. Second, the absence of a non-PCOS control group restricts interpretation to within-syndrome heterogeneity rather than disease-specific differences. Third, insulin measurements and indices such as HOMA-IR were unavailable, which may have reduced sensitivity for detecting early insulin resistance–driven phenotypes. In addition, anti-TPO antibodies exhibited higher missingness in the raw dataset, and complete-case selection may have introduced selection bias.

Anthropometric measures such as body mass index (BMI) and waist circumference were not available for the clustering feature set, which may have limited the detection of obesity-driven phenotypes and reduced the ability to disentangle adiposity-related metabolic variation.

Additionally, the cohort consisted of young hospitalized women from a single clinical center, which may limit generalizability to broader PCOS populations, including older women with more advanced metabolic complications. Medication exposure was minimized through exclusion criteria, but residual confounding cannot be fully excluded.

Despite these limitations, the study has notable strengths. We applied a transparent and clinically interpretable analytic framework combining PCA with probabilistic clustering, supported by rigorous stability validation. The integration of multiple endocrine axes together with metabolic markers represents a comprehensive approach to PCOS heterogeneity. Finally, the identification of a reproducible thyroid/autoimmune-enriched phenotype highlights the importance of endocrine comorbidity as a contributor to biologically meaningful variation within PCOS.

Taken together, our findings support a multidimensional model of PCOS heterogeneity in which thyroid autoimmunity represents a biologically meaningful axis of variation already detectable in early adulthood. Future longitudinal studies are warranted to determine whether this endocrine-autoimmune phenotype confers differential long-term cardiometabolic risk.

## Conclusions

In conclusion, our data-driven analysis identified two stable endocrine–metabolic phenotypes in young women with PCOS, including a distinct subgroup enriched for thyroid/autoimmune features. These findings demonstrate the feasibility of multidimensional phenotyping early in the disease course and suggest that thyroid-related endocrine variability may represent an underrecognized dimension of PCOS heterogeneity.

Longitudinal studies are warranted to determine whether such phenotypes predict future cardiometabolic outcomes and whether endocrine–metabolic stratification can guide personalized screening and prevention strategies in PCOS.

## DATA AVAILABILITY

The datasets generated and analyzed during the current study are not publicly available due to institutional ethical regulations and data protection policies. De-identified data may be made available from the corresponding author upon reasonable request and subject to institutional approval.

The statistical analysis code used to generate the results reported in this article is publicly available at GitHub: https://github.com/npiorkowska-science/pcos-endocrine-metabolic-phenotyping

## Clinical Trial Registration

This study was not registered as a clinical trial, as it was an observational cross-sectional analysis.

## AUTHOR CONTRIBUTIONS

NP conceptualized and designed the study.

NP, KN performed the statistical analyses, developed the analysis code, conducted the data analyses, and drafted the initial manuscript.

GF, AB, ML, KN contributed to the intellectual content of the study, critically revised the manuscript, and provided final editorial oversight.

All authors reviewed and approved the final version of the manuscript.

## ACKNOWLEDGMENTS

The authors thank the clinical and laboratory staff involved in data collection and patient care.

## SUPPLEMENTARY MATERIALS

### Supplementary Figures

Figure S1. Distributional QC and preprocessing diagnostics

Figure S2. Participant flow diagram (complete-case selection)

Figure S3A-B. PCA diagnostics

Figure S4A-D. Cluster selection metrics

Figure S5. PCA space and UMAP projection

Figure S6. Forest plot of regression

Figure S7. AMH post-hoc validation plot

### Supplementary Tables

Table S1. Feature glossary and transformations

Table S2. Missingness by feature

Table S3. PCA loading matrix

Table S4. Cluster stability metrics

Table S5. Endpoint definitions and thresholds

Table S6. Full regression output table

Table S7. AMH summary statistics by cluster

